# Efficacy of conservative physiotherapeutic approaches in hormonal-related headaches: a scoping review protocol

**DOI:** 10.1101/2022.01.10.22268916

**Authors:** Marzia Stella Yousif, Cecilia Bagnoli, Tiziano Innocenti, Paolo Bizzarri

## Abstract

**Introduction:** Headache is one of the most common and disabling conditions worldwide, as described by the World Health Organization report. The risk of suffering from headache has been described to increase from twofold to threefold in adult women compared to men, depending on the studies. These gender differences have been linked to environmental, genetic, epigenetic, and hormonal aspects. Sex hormones can enhance headaches mainly through sensitization of the trigemino-vascular system and modulation of the blood vessel factors, with significant clinical consequences. International guidelines suggest several pharmacological and non-pharmacological treatments in the management of headache disorders as acute or preventive therapies. Few studies have been conducted on the efficacy and effectiveness of therapies in managing hormonal-related headaches to date. Therefore, this scoping review (ScR) aims to summarize the evidence regarding the efficacy of conservative physiotherapeutic approaches on this topic in the domain of gender medicine, which studies sex influences on pathophysiology, clinical signs, prevention, and therapy of diseases.

**Methods and analysis:** The ScR will be performed following the 6-stage methodology suggested by Arksey and O’Malley and the extensions to the original framework recommended by the Joanna Briggs Institute. MEDLINE, Cochrane Central, Scopus, CINHAL, Embase and PEDro databases will be searched. Additional records will be identified through searching in grey literature and the reference lists of all relevant studies. No study design, publication type, language nor date restrictions will be applied. Two reviewers will independently screen all abstracts and full-text studies for inclusion. The research team will develop a data collection form to extract the studies’ characteristics. A tabular and accompanying narrative summary of the information will be provided. This protocol received input from all authors who have expertise in research methodology and specific knowledge in the field.

**Ethics and dissemination:** This study does not require ethical approval as we will not collect personal data. It will summarize information from publicly available studies in line with the nature of the study’s methodology. Regarding dissemination activities, the results of this review will be submitted for publication in a peer-reviewed journal, presented at relevant conferences in the field and disseminated through working groups, webinars and partners.

**Strengths and limitations of this study:**

- This will be the first ScR to provide a comprehensive overview of all studies dealing with conservative physiotherapeutic approaches in hormonal-related headaches.
- Results from this study will provide gender-specific and age-specific evidence of efficacy
- The results will add meaningful information for clinicians for the management of this condition and researchers to direct future research.
- A robust clinical recommendation will be possibly prevented due to limitations of the available literature.

## Introduction

Headache is one of the most common and disabling conditions worldwide, as described by the World Health Organization report. Tension-type headache (TTH) appears to be the most prevalent form of primary headache, up to 50%. At the same time, migraine is the most disabling one, particularly in female populations between 20-50 years old [1].

The risk of suffering from headache has been described to increase from twofold to threefold in women compared to men, depending on the studies[2]. Moreover, women experience higher disability, longer duration of headache attacks and recovery time, higher recurrence rate, more frequent accompanying symptoms, and are at higher risk of transition from episodic to chronic migraine[3].

These gender differences have been linked to environmental, genetic, epigenetic, and hormonal aspects. In particular, fluctuations of estrogens and progesterones can enhance headaches mainly through modulation of blood vessel factors and sensitization of the trigemino-vascular system, with significant clinical consequences. While the prevalence of headache in male and female pediatric populations is similar, menarche has often been reported as related to migraine onset. Moreover, periods of hormonal fluctuation (e.g. menses, pregnancy, peri-menopause) significantly impact headache symptoms, especially in migraineurs[4].

Although often addressed in clinical practice, the study of diagnostic procedures and treatments of hormonal-related headaches has been neglected in the literature.

The International Classification of Headache Disorders (ICHD), the reference standard for terminology and diagnostic procedures in patients suffering from headaches, includes menstrual migraine with or without aura. However, this specific diagnosis relies on headache characteristics of typical migraine attacks (e.g. pulsating quality of pain, unilateral location, photophobia, phonophobia, nausea and/or vomiting), which do not appear to apply to most people suffering from menstrual headaches that have been reported as present in up to 65% of Italian teenager samples[5]. Moreover, headache has been linked to a significant impact on women quality of life. Ishii et al. (2020)[6] described that 121 (19.9%) out of 607 migraineur women avoided pregnancy because of migraine. This population was more likely to suffer from chronic migraine or menstrual-related migraine and believed that headache would have been worse during pregnancy.

Fewer studies have been published on hormonal influences on TTH. TTH appear to be influenced to a lesser degree by pregnancy and oral contraceptive compared to migraine subjects. However, a recent study has reported that double the patients with TTH suffer from headache exclusively in the menstrual period compared to migraineurs (8% vs 4%)[7]. Based on these findings, a diagnosis of menstrual TTH has been proposed in the literature[8].

ICHD-3 refers to pregnancy-related headaches exclusively in specific disorders, such as *12.1 Headache attributed to somatization disorder*, occurring as part of the symptomatic presentation of a somatization disorder of the psychic sphere, or *10.3.4 Headache attributed to pre-eclampsia or eclampsia*. No mention is made to menopause factors.

International guidelines suggest several pharmacological and non-pharmacological treatments in the management of headache disorders as acute or preventive therapies. Few studies have been conducted on the efficacy and effectiveness of therapies in managing hormonal-related headaches to date.

Headaches are multifactorial neurological conditions influenced by mood, diet, sleep quality, painful and non-painful comorbidities, and physical activity. Conversely, approaches based on these premises can impact modulating clinical presentations. For example, exercise therapy has shown promising results in modulating estrogen and progesterone levels in perimenopausal women[9], such as manual therapy in managing primary headaches[10]. However, a review on the efficacy of rehabilitation approaches on hormonal-related headaches is still missing.

Therefore, this scoping review (ScR) aims to summarize the evidence regarding the efficacy of conservative physiotherapeutic approaches on this topic in the domain of gender medicine, which studies sex influences on pathophysiology, clinical signs, prevention, and therapy of diseases[11]. As maintained by the Joanna Briggs Institute (JBI), the ScR approach may be used to map and clarify key concepts, identify gaps in the research knowledge base, examine how research is conducted on a specific topic or field and inform future research[12]. For this reason, other types of review, such as systematic reviews, umbrella reviews or rapid reviews, were not deemed methodologically effective.

## Methods and analysis

The proposed ScR will be performed following the 6-stage methodology suggested by Arksey and O’Malley (2005)[13] and conducted following the extensions to the original framework recommended by the Joanna Briggs Institute methodology (JBI) for ScR[12].

The 6-stage are outlined as follows: (1) identification of the research question, (2) identification of relevant studies, (3) selection of studies, (4) charting of data, (5) summary of results and (6) consultation with stakeholders, an optional but recommended evaluation component of a ScR.

The Preferred Reporting Items for Systematic reviews and Meta-Analyses extension for Scoping Reviews (PRISMA-ScR) Checklist for reporting will be used to report the final manuscript[14]. We will follow the framework of Population, Concept and Context (PCC) proposed by The Joanna Briggs Institute to describe the elements of the inclusion criteria.

### Stage 1: Identifying the research question

We formulated the following research question: “What is known from the existing literature about the evidence regarding the efficacy of conservative physiotherapeutic approaches in hormonal-related headaches?”

In particular, the objectives of this study will be to:

1. Provide a comprehensive overview of all studies dealing with conservative physiotherapeutic approaches in hormonal-related headaches.
2. Identify and analyze the terminology used in the included studies to describe these conditions and other conditions that could emerge from the analysis of the included studies.
3. Identify any knowledge gaps in the physiotherapeutic approach of hormonal-related headaches.

### Stage 2: Identifying relevant studies

All named authors have participated in an iterative process to develop the initial search strategy. This includes identifying key terms, inclusion and exclusion criteria and relevant databases.

#### Inclusion criteria

In a ScR, the three elements of population, concept and context are used to establish inclusion criteria[12]. The population details the relevant characteristics of participants, the concept is the principal focus of the review, and the context describes the setting under examination.

#### Population

This review will consider studies that include women with headaches related to their menstrual cycle as a primary criterion, from menarche to the menopausal stage, including pregnant women. The menopausal stage is defined as the permanent cessation of menses (menstruation), usually determined after 6 to 12 months of amenorrhea.

#### Concept

This review will include studies that explore and report the efficacy of any conservative physiotherapeutic approaches in hormonal-related headaches.

#### Context

This review will consider studies conducted in any context. Studies that do not meet the above stated Population-Concept-Context (PCC) criteria or provide insufficient information will be excluded.

#### Types of evidence sources

For the purposes of this ScR, we will include evidence from any publication type, including (but not limited to) primary research studies (quantitative, qualitative and mixed-methods), systematic reviews, meta-analyses, guideline implementation. No restriction regarding study design, time, location, language and setting will be applied.

#### Search strategy

The research group will develop a three-step approach.

1. A preliminary search in PubMed was undertaken to identify articles on the topic. Due to the lack of shared classifications or terminology, we analyzed all the terms regarding hormonal factors in headache patients, terms included in the titles and abstracts of relevant articles, and the index terms Table 1. Variants of the terms identified in Table 1 were refined to create a second search strategy with free text words and Medical Subject Headings (MeSH) terms. The information gained from the initial search was used to develop a more comprehensive search strategy based on the PCC framework for PubMed. Table 1 shows the initial search strategy to be executed.
2. A final comprehensive search will be conducted across MEDLINE (through PubMed interface), Cochrane Central, Scopus, CINHAL, Embase, and PEDro. The search strategy will be adapted in the database mentioned above, including all identified keywords and index terms.
3. In addition, also grey literature (e.g. Google Scholar) and the reference lists of included studies will be searched manually to identify any additional studies that may be relevant for this review. The PRISMA-S will be used to report the search strategies[15].

### Stage 3: study selection

The search results will be collated and exported to EndNote V.X9 (Clarivate Analytics, PA, USA). Duplicates will be automatically removed before the file containing a set of unique records is made available to reviewers for further processing (i.e. study screening and selection). The review process will consist of two levels of screening using Rayyan QCRI online software[16]: (1) a title and abstract review and (2) a full-text review. Two investigators will screen the articles independently for both levels to determine if they meet the inclusion/exclusion criteria. In case of any disagreement on inclusion, both reviewers will review full-text articles again. If an agreement cannot be reached, this will be resolved by an independent third reviewer adjudication. Reasons for the exclusion of any full-text source of evidence will be recorded and reported in the ScR report.

**Table 1.**
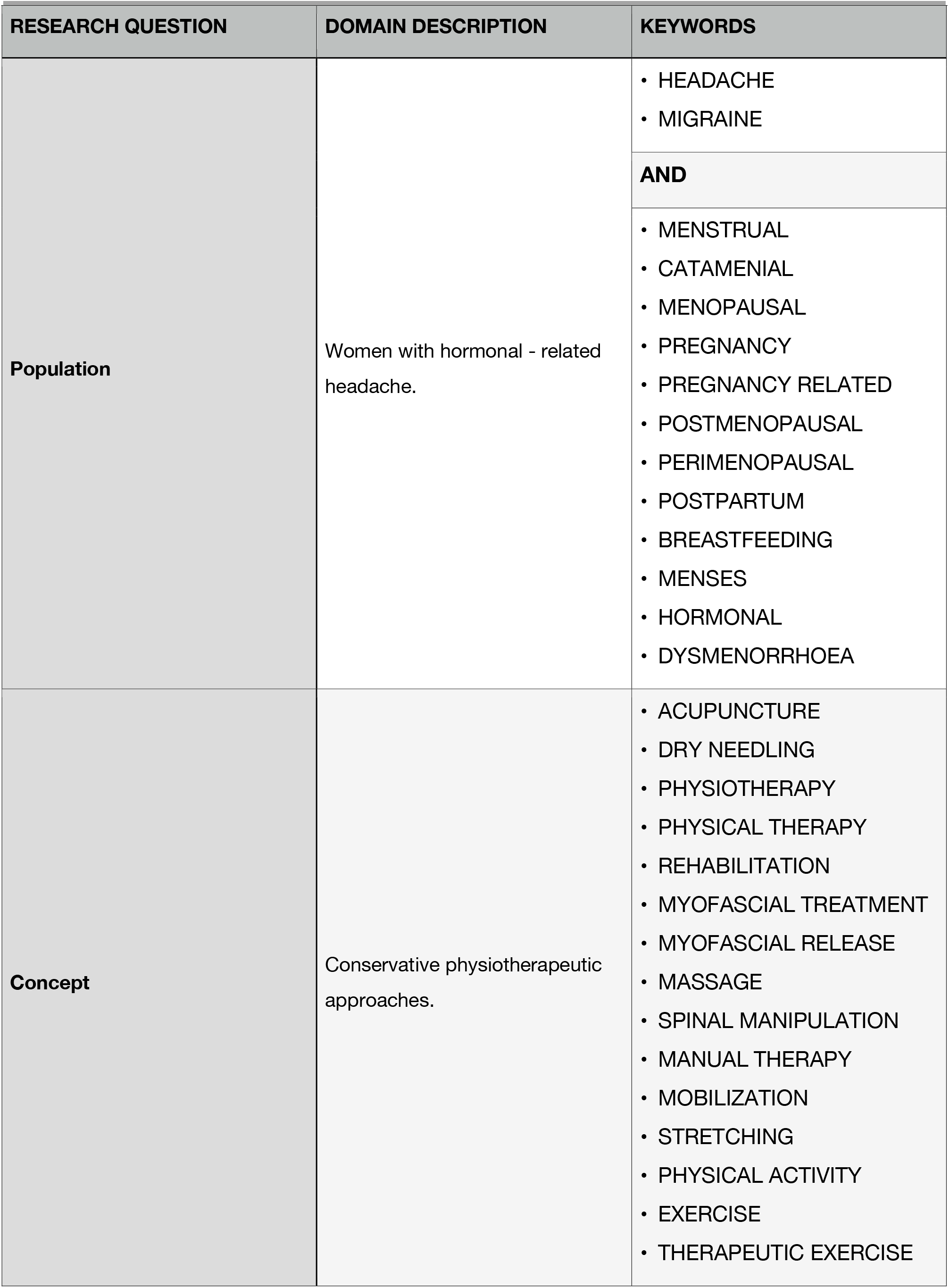
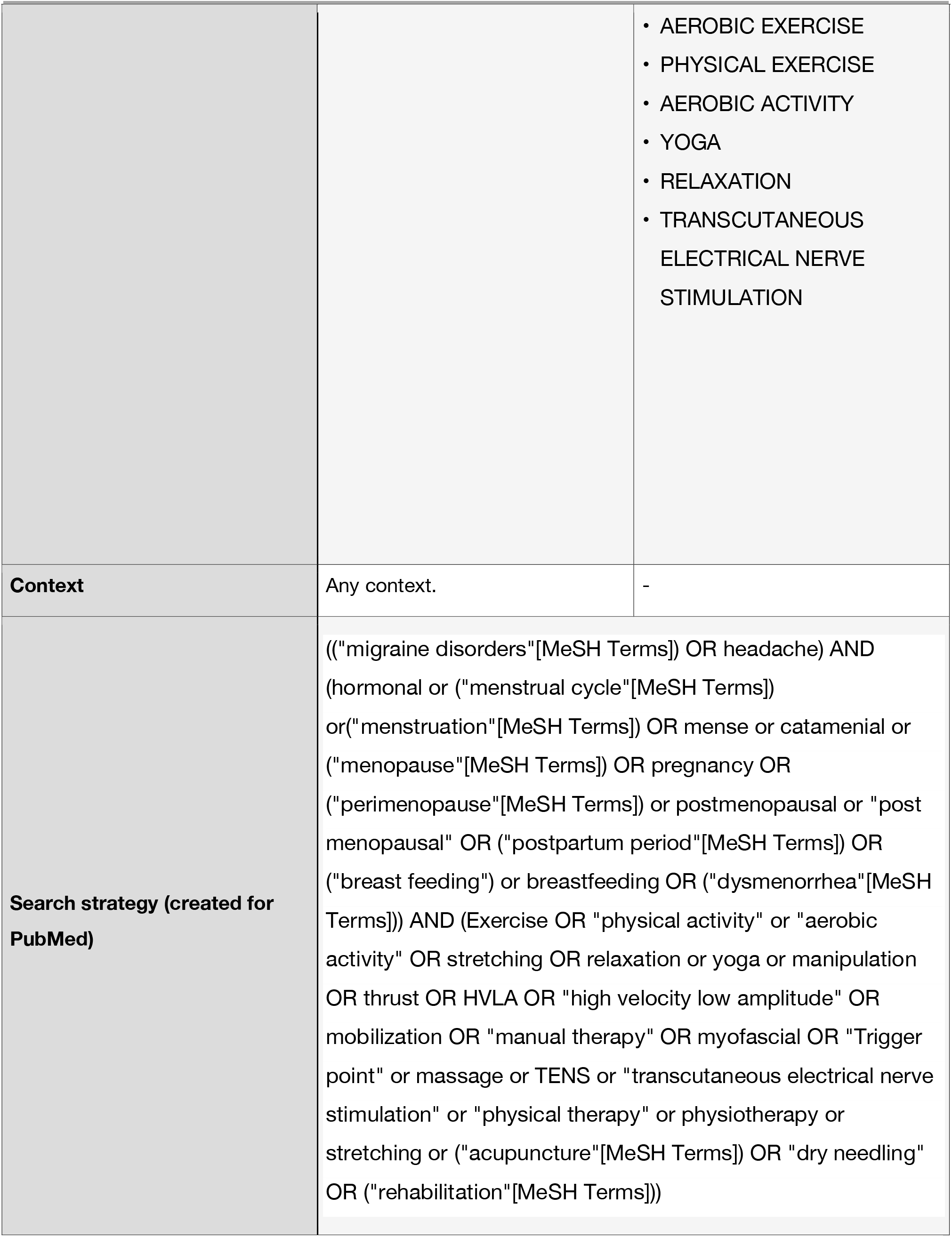
Search strategy.

The results of the study selection will be reported in full in the final manuscript and summarised in a Preferred Reporting Items for Systematic Reviews and Meta-analyses (PRISMA) flow diagram[17]. (Figure 1).

**Figure 1.**
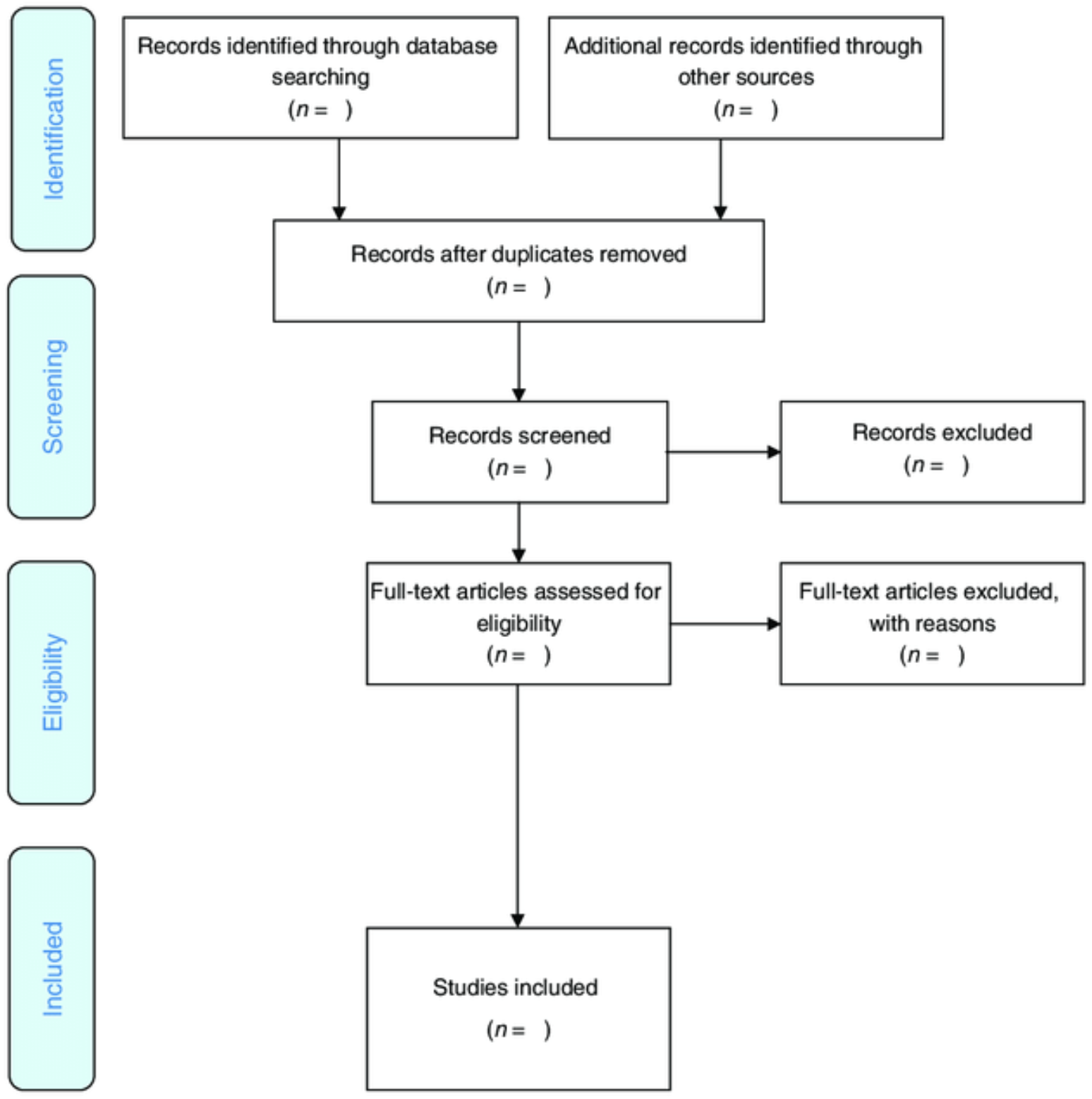

### Stage 4: charting the data

The research team will develop a data collection form to extract the characteristics of the included studies. The final sample of articles will be divided equally among the reviewers, with two researchers reviewing each article. The extraction tool will be used to collect and chart data. The two reviewers will meet to resolve conflicts, hone their shared understanding of the extraction method and refine the tool when needed. A third researcher will resolve unreconciled disputes. Any modifications to the data extraction strategy will be reported in the results section of the final ScR.

The initial data-collection form will include the following elements:

- Author(s), year of publication, study location
- Intervention type and comparator (if any); duration of the intervention
- Study populations
- Aims of the study
- Methodology
- Outcome measures
- Important results

### Stage 5: Collating, summarizing and reporting the results

Results from the ScR will be summarized descriptively through tables and diagrams. Specifically, we will summarize and synthesize the data extracted from the studies and highlighting any knowledge and implementation gaps for priority populations.

The objectives of each selected study, the concepts or approaches adopted in each article, and the results related to the study research question will be summarized and explained in the results.

### Stage 6: consultation with stakeholders

This will be the last step of the review. After analyzing and interpreting, preliminary results will be presented to a group of clinical experts. This strategy aims to share preliminary study findings (knowledge transfer and exchange), obtain potentially relevant studies not included in the initial search and develop effective dissemination strategies and directions for future studies[18].

#### Patient and public involvement

This protocol was developed without patient involvement. Patients were not invited to comment on the protocol design and were not consulted to synthesize outcomes or interpret the results. Patients were not invited to contribute to the writing or editing of this document for readability or accuracy. The results of this ScR will inform the development and design of a research study for which patient and public partnership will be sought.

#### Ethics and dissemination

This study does not require ethical approval as we will not collect personal data. It will summarize information from publicly available studies in line with the nature of the study’s methodology. Regarding dissemination activities, the results of this review will be submitted for publication in a peer-reviewed journal, presented at relevant conferences in the field and disseminated through working groups, webinars and partners. The researchers also intend to form recommendations for areas of future research.

Headache has been linked to a significant impact on women quality of life[6]. The ScR of the efficacy of conservative physiotherapeutic approaches in hormonal-related headaches in this protocol will survey the current scientific literature to identify gaps in the research across contents and methods to encourage future research and uniformity of terms and prevent duplication of efforts.

## Data Availability

All data produced in the present study are available upon reasonable request to the authors

## Contributors

All authors designed the protocol, reviewed the manuscript, approved the final version and participated in the 6-stage.

## References

1 Stovner LJ, Nichols E, Steiner TJ, et al. Global, regional, and national burden of migraine and tension-type headache, 1990–2016: a systematic analysis for the Global Burden of Disease Study 2016. Lancet Neurol 2018;17:954–76. doi:10.1016/S1474-4422(18)30322-3

2 Todd C, Lagman-Bartolome AM, Lay C. Women and Migraine: the Role of Hormones. Curr Neurol Neurosci Rep 2018;18:42. doi:10.1007/s11910-018-0845-3

3 Allais G, Chiarle G, Sinigaglia S, et al. Gender-related differences in migraine. Neurol Sci Off J Ital Neurol Soc Ital Soc Clin Neurophysiol 2020;41:429–36. doi:10.1007/s10072-020-04643-8

4 Buse DC, Loder EW, Gorman JA, et al. Sex differences in the prevalence, symptoms, and associated features of migraine, probable migraine and other severe headache: results of the American Migraine Prevalence and Prevention (AMPP) Study. Headache 2013;53:1278–99. doi:10.1111/head.12150

5 Bianchin L, Bozzola M, Battistella Pier A, et al. Menstrual Cycle and Headache in Teenagers. Indian J Pediatr 2019;86:25–33. doi:10.1007/s12098-018-2829-3

6 Ishii R, Schwedt TJ, Kim S-K, et al. Effect of Migraine on Pregnancy Planning: Insights From the American Registry for Migraine Research. Mayo Clin Proc 2020;95:2079–89. doi:10.1016/j.mayocp.2020.06.053

7 Karlı N, Baykan B, Ertaş M, et al. Impact of sex hormonal changes on tension-type headache and migraine: a cross-sectional population-based survey in 2,600 women. J Headache Pain 2012;13:557–65. doi:10.1007/s10194-012-0475-0

8 Arjona A, Rubi-Callejon J, Guardado-Santervas P, et al. Menstrual tension-type headache: evidence for its existence. Headache 2007;47:100–3. doi:10.1111/j.1526-4610.2007.00656.x

9 Kossman DA, Williams NI, Domchek SM, et al. Exercise lowers estrogen and progesterone levels in premenopausal women at high risk of breast cancer. J Appl Physiol 2011;111:1687–93. doi:10.1152/japplphysiol.00319.2011

10 Falsiroli Maistrello L, Geri T, Gianola S, et al. Effectiveness of Trigger Point Manual Treatment on the Frequency, Intensity, and Duration of Attacks in Primary Headaches: A Systematic Review and Meta-Analysis of Randomized Controlled Trials. Front Neurol 2018;9:254. doi:10.3389/fneur.2018.00254

11 Baggio G, Corsini A, Floreani A, et al. Gender medicine: a task for the third millennium. Clin Chem Lab Med 2013;51:713–27. doi:10.1515/cclm-2012-0849

12 Peters MDJ, Godfrey C, McInerney P, Munn Z, Tricco AC, Khalil, H. Chapter 11: Scoping Reviews (2020 version). In: Aromataris E, Munn Z (Editors). JBI Manual for Evidence Synthesis, JBI, 2020. Available from https://synthesismanual.jbi.global. https://doi.org/10.46658/JBIMES-20-12.

13 Arksey H, O’Malley L. Scoping studies: towards a methodological framework. Int J Soc Res Methodol 2005;8:19–32. doi:10.1080/1364557032000119616

14 Tricco AC, Lillie E, Zarin W, et al. PRISMA Extension for Scoping Reviews (PRISMA-ScR): Checklist and Explanation. Ann Intern Med 2018;169:467–73. doi:10.7326/M18-0850

15 Rethlefsen ML, Kirtley S, Waffenschmidt S, et al. PRISMA-S: an extension to the PRISMA Statement for Reporting Literature Searches in Systematic Reviews. Syst Rev 2021;10:39. doi:10.1186/s13643-020-01542-z

16 Ouzzani M, Hammady H, Fedorowicz Z, et al. Rayyan-a web and mobile app for systematic reviews. Syst Rev 2016;5:210. doi:10.1186/s13643-016-0384-4

17 Moher D, Liberati A, Tetzlaff J, et al. Preferred reporting items for systematic reviews and meta-analyses: the PRISMA statement. Ann Intern Med 2009;151:264–9, W64. doi:10.7326/0003-4819-151-4-200908180-00135

18 Levac D, Colquhoun H, O’Brien KK. Scoping studies: advancing the methodology. Implement Sci IS 2010;5:69. doi:10.1186/1748-5908-5-69

